# Sleep and circadian indices for planning post-pandemic university timetables

**DOI:** 10.1101/2022.01.05.22268660

**Authors:** Sara Montagnese, Lisa Zarantonello, Chiara Formentin, Gianluca Giusti, Chiara Mangini, Cheryl M. Isherwood, Paolo Ferrari, Antonio Paoli, Daniela Mapelli, Rosario Rizzuto, Stefano Toppo, Debra J. Skene, Roberto Vettor, Rodolfo Costa

**Affiliations:** Department of Medicine, University of Padova, Padova, Italy; Chronobiology Section, Faculty of Health and Medical Sciences, University of Surrey, Guildford, UK; reMedia Web Agency S.r.l., Padova, Italy; Department of Biomedical Sciences, University of Padova, Padova, Italy; Department of General Psychology, University of Padova, Padova, Italy; Department of Molecular Medicine, University of Padova, Padova, Italy; Department of Biology, University of Padova, Padova, Italy; Institute of Neuroscience, National Research Council (CNR), Padova, Italy

**Keywords:** circadian hygiene, chronotype, sleep, university students, academic performance, pandemic

## Abstract

The aims of the present study were to obtain sleep quality and sleep timing information in a group of university students, and to evaluate the effects of a circadian hygiene education initiative. All students of the University of Padova (approximately 64,000) were contacted by e-mail (major campaigns in October 2019 and October 2020) and directed to an *ad hoc* website for collection of demographics and sleep quality/timing information. Participants (n=5740) received one of two sets of circadian hygiene advice (*“A regular life”* or *“Bright days and dark nights”*). Every month, they were then asked how easy it had been to comply, and provided with the advice again. At any even month from joining, they completed the sleep quality/timing questionnaires again. Information on academic performance was obtained *post hoc*, together with representative samples of lecture (n=5972) and exam (n=1800) timings, plus lecture attendances (n=25,302). 52% of students had poor sleep quality and 82% showed signs of sleep deprivation. Those who joined in October 2020, after several months of lockdown and distance learning, had better sleep quality, less sleep deprivation and later sleep habits. The *“Bright days and dark nights”* advice resulted in earlier get-up time/midsleep compared to the *“A regular life”* advice. Significant changes in most sleep quality and sleep timing variables were observed in both advice groups over time, also in relation to pandemic-related events characterising 2020. Early-chronotype students had better academic performances compared to their later chronotype counterparts. In a multivariate model, sleep quality, chronotype and study subject were independent predictors of academic performance. Taken together, these results underlie the importance of designing circadian-friendly university timetables.

## Introduction

Chronotype reflects an individual’s natural inclination to place their activity/sleep within different intervals of the 24-hour day, and is partly genetically determined (Ashbrook et al., 2020). Children tend towards earlier, morning chronotypes, whereas adolescents and young adults exhibit a sharp change towards eveningness (which is more pronounced/prolonged in males), but then mature adults/older individuals become progressively more morning-like (Roenneberg et al., 2004). Late-chronotype individuals are penalised by the Western “social clock” which forces them to study/work/function during the early part of the day, counter to their natural biological clock (Gentry et al., 2021). It has been reported that early classes/early school tests in evening-type adolescents adversely affect their academic performance, especially in scientific subjects (Zerbini et al., 2017), and that changes in school start times may help improve sleep (Winnebeck et al., 2020; Meltzer et al., 2021) and, possibly, also grades (Zerbini et al., 2017). Less but similar information is available on the relationship between study timetables, sleep timing/length and academic performance in university students (Li et al., 2018, Beşoluk et al., 2011; Smarr and Schirmer, 2018). In addition, there are also studies on the effects of sleep deprivation on university students’ athletic performance and wellbeing (Hodge et al., 2012; Bolin, 2019).

The aims of this study were to obtain comprehensive sleep quality/timing information, and to evaluate the effects of a university welfare-based circadian hygiene initiative - directed at all students - on sleep and academic performance. As the study progressed, it became apparent that novel information could also be obtained on the effects of the pandemic-related lockdown on sleep-wake behaviour.

## Methods

### Overview of the survey and educational initiative

An *ad hoc* full-responsive website was created for collection of the relevant information, with either an Italian or English interface to choose from. All active students at the University of Padova (approximately 64,000) were contacted by the university welfare offices on 28 October 2019 with an e-mail message including a brief description of the initiative and its aims, an invitation to participate, and a “call to action” button leading to the website (https://sleeprhythm-unipd.it/). This could only be accessed by use of the personal Shibboleth University of Padova username/password. Students who joined the initiative went on to receive reminders on the 28^th^ of each subsequent month to access the website again to provide additional information and receive advice. The website was always open for newcomers to join; access after each reminder was possible for 7 days. On joining for the first time, students were asked to provide information on demographics, sleep-wake timing, night sleep quality and diurnal sleepiness. They then received one of two sets of circadian hygiene advice with similar, expected effects *(“A regular life”* or *“Bright days and dark nights”*, Figure 1). Every month after joining, they were asked how easy it had been to comply with the advice and were provided with the advice again. At every even month from joining, they were asked to complete demographic, sleep-wake timing, night sleep quality and diurnal sleepiness information again. They were also asked how easy it had been to comply, and finally, they were provided with the advice again (Supplementary Table 1). An additional information/recruitment campaign was run on 28 October 2020 that excluded any student who had already joined. Information on the participating students’ career parameters was obtained from the career office *post hoc*. Representative samples of the timings of electronic room bookings for lectures/exams and of recorded, anonymous lecture attendances were also obtained *post hoc*.

**Figure 1.**
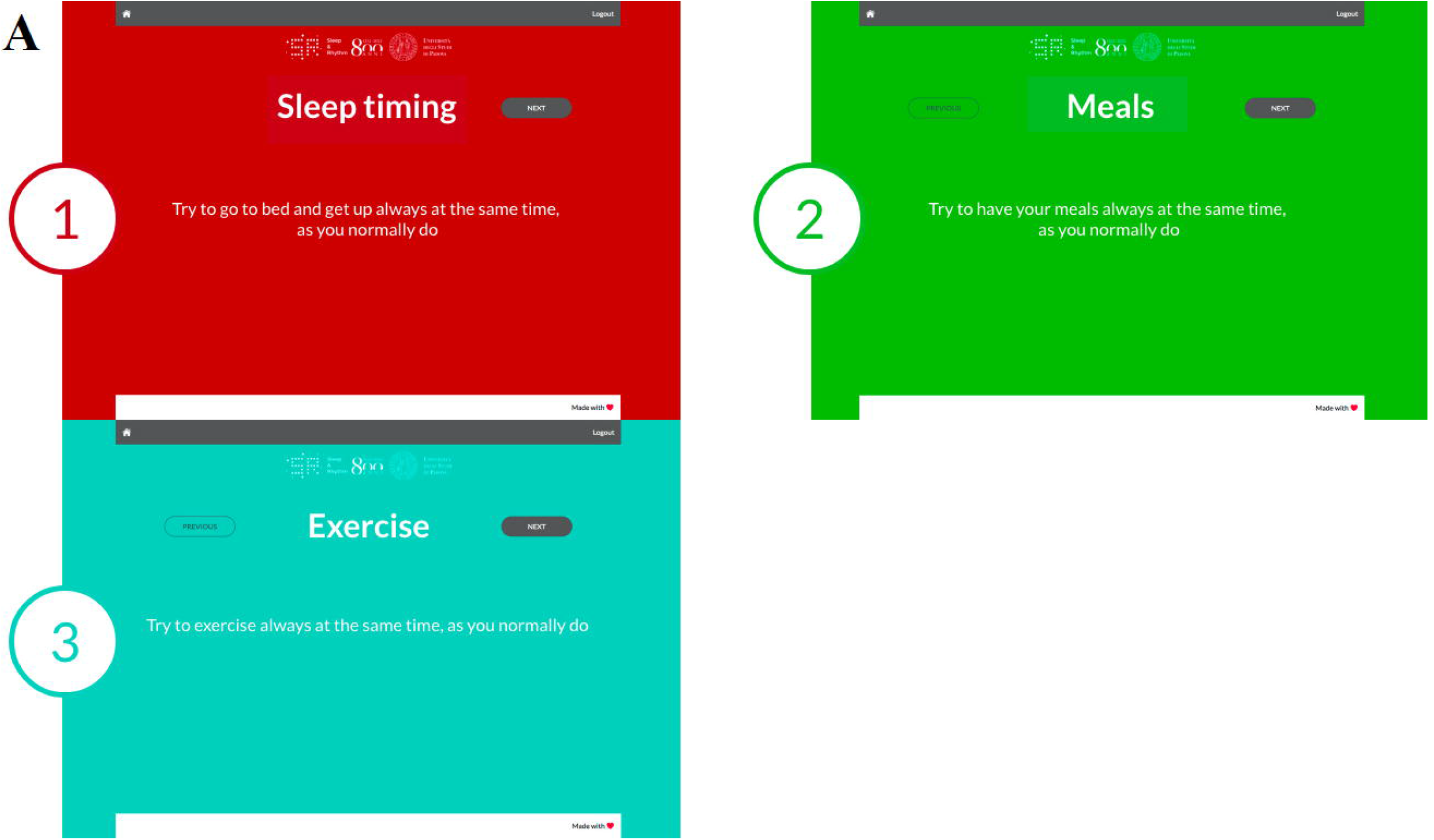
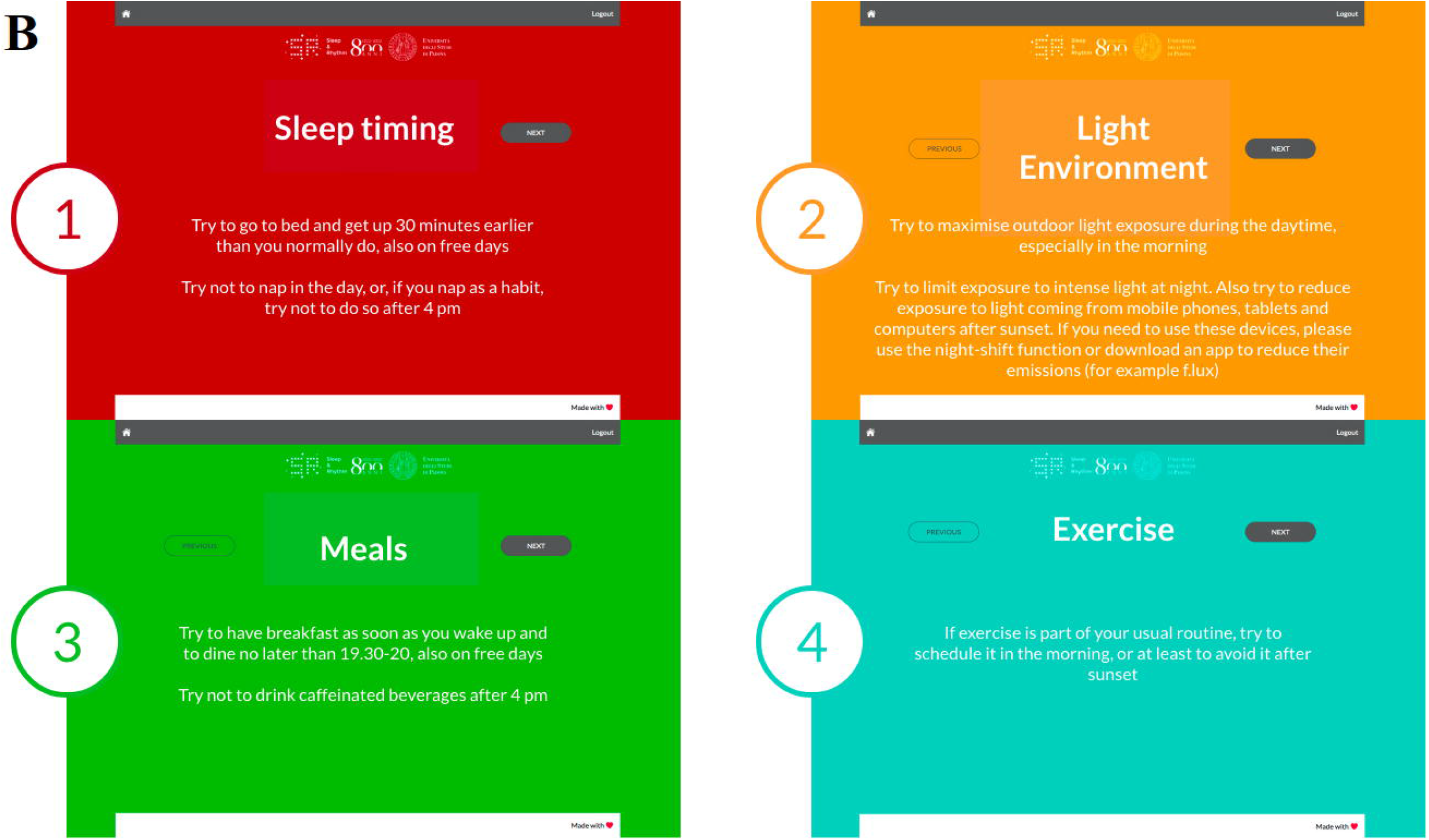
Circadian hygiene advice. Advice provided in the *“A regular life”* (**A**) and the *“Bright days and dark nights”* (**B**) groups. The numbering of each panel refers to the temporal sequence in which the advice was provided (on subsequent smart phone/tablet/computer screens).

### Demographics

Date and time of both first accessing and completing all procedures scheduled at time 0 was obtained. In addition to basic demographics, students also provided information on their height, weight, any problems with eyesight, any disease and/or any medication they might be on. In relation to eyesight, diseases and medication, a yes/no answer was a requirement in order to proceed. By contrast, the field to provide additional details on any of these three items could be left empty. Authors SM, CF and CM, who are all practicing clinicians, independently classified listed eyesight problems, diseases and drugs as worth considering or negligible; where their classifications differed, agreement was sought for and reached.

### Sleep-wake profile

Carefully developed, user-friendly electronic versions with equal resolution and ease of completion on mobile phones, tablets and computers of the following questionnaires were then presented on the full-responsive website:

#### The Sleep Timing Sleep Quality Screening (STSQS) questionnaire

(Montagnese et al., 2009). This provides a simple, overall assessment of sleep quality rated on a 0-10 visual-analogue scale (0 = worst, 10 = best sleep ever) and allows collection of information on habitual sleep timing (i.e. bed time, try to sleep time, sleep latency, wake up and get up time).

#### The Pittsburgh Sleep Quality Index (PSQI)

(Buysse et al., 1989, Curcio et al., 2013). Responses to 19 questions are used to generate seven components (subjective sleep quality, sleep latency, sleep duration, sleep efficiency, sleep disturbance, use of sleep medication, daytime dysfunction), each of which is scored from zero (best) to three (worst). These component scores are then summated to provide the total PSQI score (range: 0-21); scores of >5 identify ‘poor sleepers’ (Buysse et al., 1989).

#### The Epworth Sleepiness Scale (ESS)

(Johns, 1991; Vignatelli et al., 2003). Subjects rate their likelihood of ‘dozing off’ in eight different day-time situations, on a scale of zero (unlikely) to three (very likely). The component scores are summated to provide a total score (range: 0-24); a score of ≥11 is considered abnormal (Johns, 1991).

#### The self-morningness/eveningness (Self-ME) questionnaire

(Turco et al., 2015). This is a validated, single-question assessment of chronotype through which participants qualify themselves as definitely morning, morning, evening or definitely evening types.

#### The ultra-Short version of the Munich ChronoType Questionnaire (µMCTQ)

(Ghotbi et al., 2020). This is an adaptation of the MCTQ (Roenneberg et al., 2003) from 17 to 6 essential questions, allowing for a quick assessment of midsleep (i.e. the midpoint, expressed as clock time, between sleep onset and sleep offset), social jetlag (in this instance, the uncorrected difference between midsleep on free and work/study days) and sleep duration. Social jetlag was both treated as a continuum variable and also qualified as *positive* (difference between midsleep on free and work/study days > +20 min), *absent* (−20 min < difference < +20 min) and *negative* (difference < - 20 min) (Korman et al., 2020).

### Circadian hygiene advice and compliance

Two alternative sets of advice (novel in their formulation but grounded in circadian practice; Abbot et al., 2015) were provided (and remained the same on every subsequent “refresh” monthly reminder) with similar, expected effects exerted via two different forms of habits’ adjustment (Facer-Childs et al., 2019). Both included elements of the recommendations routinely provided to patients with either a formal diagnosis and/or features of delayed sleep phase type (Abbot et al., 2015), which is common in adolescents and young adults.

***“A regular life”***, encouraging participants to regularize habits in relation to sleep-wake timing, meals, and exercise (Figure 1A). This intervention did not include advice on light exposure, as while the idea of sleep, meals and exercise at regular times is extremely intuitive and easily interpreted, that of “light at regular times” seems less so. Therefore, when planning the intervention, we reasoned that advice on “light at regular times” would be prone to misinterpretation.

***“Bright days and dark nights”***, encouraging participants to advance their sleep-wake, meals, and exercise timing, and to maximise/minimise light exposure in the first/second part of the day, respectively (Figure 1B).

The user-friendly, short format of the interventions was chosen to maximise the likelihood of them being read and considered by young, healthy individuals on a monthly basis, and most likely on the screen of their mobile phone. Along the same lines, we did not modulate advice in relation to photoperiod or daylight saving time (DST), deliberately opting for repeated, identical advice, which could be read, re-read (on refresh) and memorised easily, hopefully becoming part of the students’ routine.

This being a University-based welfare initiative all students were provided with advice, i.e. none served as “placebo”.

#### Compliance

Participants subjectively rated their compliance with each piece of advice received on a 0 (never) to 10 (all the time) visual-analogue scale.

### Academic performance

Information was obtained from the student career offices about each student’s university course, course year, total number of exams passed, total number of “credits” (i.e. packages of formal lectures/practicals of variable length, depending on study subjects) and average marks (0-30, 18= pass) at fixed dates of the year, on a three-monthly interval (28 October 2019, 28 January 2020, 28 April 2020, 28 July 2020 and 28 October 2020). These data were then matched with the “join the initiative” date, and organised as baseline (T0) and subsequent time intervals. Studies were classed as health&medical (M), science&technology (S) and social&humanities (H), based on Italian Ministry of University and Research reference tables. Courses have variable duration: most have a three plus two year structure (in this instance students who went on to attend their master’s were qualified as fourth or fifth year), while a minority have a four to six year duration. Based on the information they provided and on University of Padova criteria, students were classified as on-site, commuters or off-site (i.e. non-compulsory lectures and/or registered for exams only).

Significant differences in sleep-wake and other parameters were observed between students pursuing M, S and H studies, which were thought to depend, to some extent, on lectures timing/duration and/or exams timing. Thus, a sample of electronic room bookings for lectures (n=5972) was obtained *post hoc* on a representative week (4-10 November 2019, i.e. close to study start; lectures were arbitrarily qualified as *morning/afternoon* if they started prior to/later than 13:00 h, respectively) and a sample of anonymous students’ recorded attendances (electronic swipe in - swipe out, n=25,302; multiple swipe ins - swipe outs, n=7765) on 30 October 2020. This date was chosen because, due to novel, pandemic-related rules, all students were asked to swipe in - swipe out electronically. Finally, a sample of exam booking times was obtained, on a representative week [9-13 September 2019 (n=1800 bookings)]; again, exams were arbitrarily qualified as *morning/afternoon* if they started prior to/later than 13:00 h, respectively.

### Study approval

The initiative was approved by the University of Padova (16 July 2019 board meeting). Students were asked to accept/tick a general data protection regulation (GDPR)-compliant informed consent, including the wording “data may be used anonymously, in aggregate fashion for scientific purposes”. The study plan was then approved by the local ethics committee.

### Statistical Analysis

Descriptive results are expressed as mean±SD/SE or as count/percentage. Normality was tested for by the Shapiro-Wilk’s test. Differences between normally distributed variables were examined by Student’s t test/one way ANOVA (*post hoc*: Scheffé test). Differences between non-normally distributed variables were examined by Mann-Whitney U test/Kruskal-Wallis ANOVA (*post hoc*: median test). Bonferroni correction for multiple comparisons was applied as appropriate. The effects of the circadian hygiene initiative and academic performance over time were analysed by repeated measures ANOVA (no missing data imputation procedures applied); η _p_^2^ was utilised as an indicator of the effect size. Factorial ANOVA was utilised for multiple grouping factors. A linear regression model was utilised to identify independent predictors of academic performance. Analyses were carried out with Statistica, version 13.1 (Dell, Round Rock, Texas, USA) and subsequently version 14.0.0.15 (TIBCO, Palo Alto, California, USA).

## Results

On 19 December 2020, 5740 students (38% males; age 23±5 yrs) had completed the first full set of questions; 55% had responded to the October 2019 call, 29% to the October 2020 call and 16% had joined at different times between October 2019 and December 2020. 740 (31% males) students attended health&medical (M), 2439 (58% males) science&technology (S) and 2492 (22% males) social&humanities (H) studies; 69 attended single courses. 2981 students were assigned to the *“A regular life”* and 2759 to the *“Bright days and dark nights”* advice group. 1144 students had one/more diseases and 1359 were on medication, with significant overlap (χ^2^=869, p<0.0001); 1295 reported minor eyesight issues.

### Sleep-wake profiles

2972 students had an abnormal Pittsburgh Sleep Quality Index (PSQI), with the most heavily affected components being subjective sleep quality, sleep latency, sleep disturbance and daytime dysfunction; 714 had an abnormal Epworth Sleepiness Scale (ESS). Based on the self-morningness/eveningness questionnaire (Self-ME), 570 students qualified themselves as definitely morning, 1896 as morning, 2233 as evening and 1041 as definitely evening. Based on the ultra-short Munich ChronoType Questionnaire (µMCTQ), 240 had negative, 755 no and 4703 positive social jetlag; 42 did not provide sufficient information. Average midsleep on study and free days was 03:54±01:06 and 05:06±01:12 (hh:mm), respectively. Average sleep duration on study and free days was 7.5±1.2 and 8.4±1.2 h, respectively.

Female students had earlier sleep-wake timing habits, more night awakenings, worse sleep quality and more daytime sleepiness compared to male students (Table 1). In addition, they had earlier midsleep on both study and free days, and they more commonly classified themselves as morning chronotypes. Finally, their sleep duration was longer on free days (Table 1). Commuters had earlier sleep-wake timing habits (e.g. bed time: 23:24±01:00 *vs*. 23:30±01:00 hh:mm, p<0.001) compared to on-site students. Students attending H were older than those attending M who, in turn, were older than those attending S studies (Table 2). Students attending M had earlier sleep-wake timing compared to students attending S/H studies. Finally, students attending H had worse sleep quality than those attending M/S studies (Table 2).

**Table 1.** Sleep-wake indices (mean±SD) on joining the initiative, by sex

| Questionnaire | Variable | Females<br>(n=3566) | Males<br>(n=2174) |
| --- | --- | --- | --- |
| **STSQS** | Bed time (hh:mm) | 23:18±00:54 | 23:42±01:06* |
|  | Try to sleep time (hh:mm) | 23:51±00:57 | 24:06±01:04* |
|  | Sleep latency (min) | 24±24 | 20±18* |
|  | Calculated sleep onset time (hh:mm) | 24:15±01:06 | 24:26±01:09* |
|  | Night awakenings (n) | 1.0±1.2 | 0.8±1.0* |
|  | Wake-up time (hh:mm) | 07:34±01:09 | 07:39±01:15 |
|  | Get-up time (hh:mm) | 07:56±01:16 | 08:00±01:21 |
|  | Latency between wake-up and get-up time (min) | 21±28 | 20±31 |
| **PSQI** | Total score (0-21) | 6.5±3.2 | 5.8±2* |
| **ESS** | Total score (0-24) | 6.7±3.6 | 5.7±3.3* |
| **Self-ME** | Definitely morning/morning/evening/definitely evening (%) | 11/34/38/16 | 8/31/40/21* |
| **μMCTQ** | Sleep onset study days (hh:mm) | 24:03±01:14 | 24:15±01:15* |
|  | Sleep offset study days (hh:mm) | 07:34±01:17 | 07:42±01:21* |
|  | Sleep duration study days (hours) | 7.5±1.2 | 7.4±1.1 |
|  | Midsleep study days (hh:mm) | 03:49±01:05 | 04:00±01:09* |
|  | Sleep onset free days (hh:mm) | 24:27±01:15 | 01:06±01:20* |
|  | Sleep offset free days (hh:mm) | 09:14±01:21 | 09:24±01:29* |
|  | Sleep duration free days (hours) | 8.4±1.2 | 8.3±1.1* |
|  | Midsleep free days (hh:mm) | 05:00±01:09 | 05:15±01:16* |
|  | Social jet-lag (hours) | 1.2±1.3 | 1.3±1.4 |

\* $p < 0.001$ (Bonferroni-corrected)
STSQS: Sleep Timing Sleep Quality Screening questionnaire; PSQI: Pittsburgh Sleep Quality Index
ESS: Epworth Sleepiness Scale; Self-ME: Self-morningness/eveningness questionnaire; μMCTQ: Ultra-Short Munich ChronoType Questionnaire

**Table 2.**
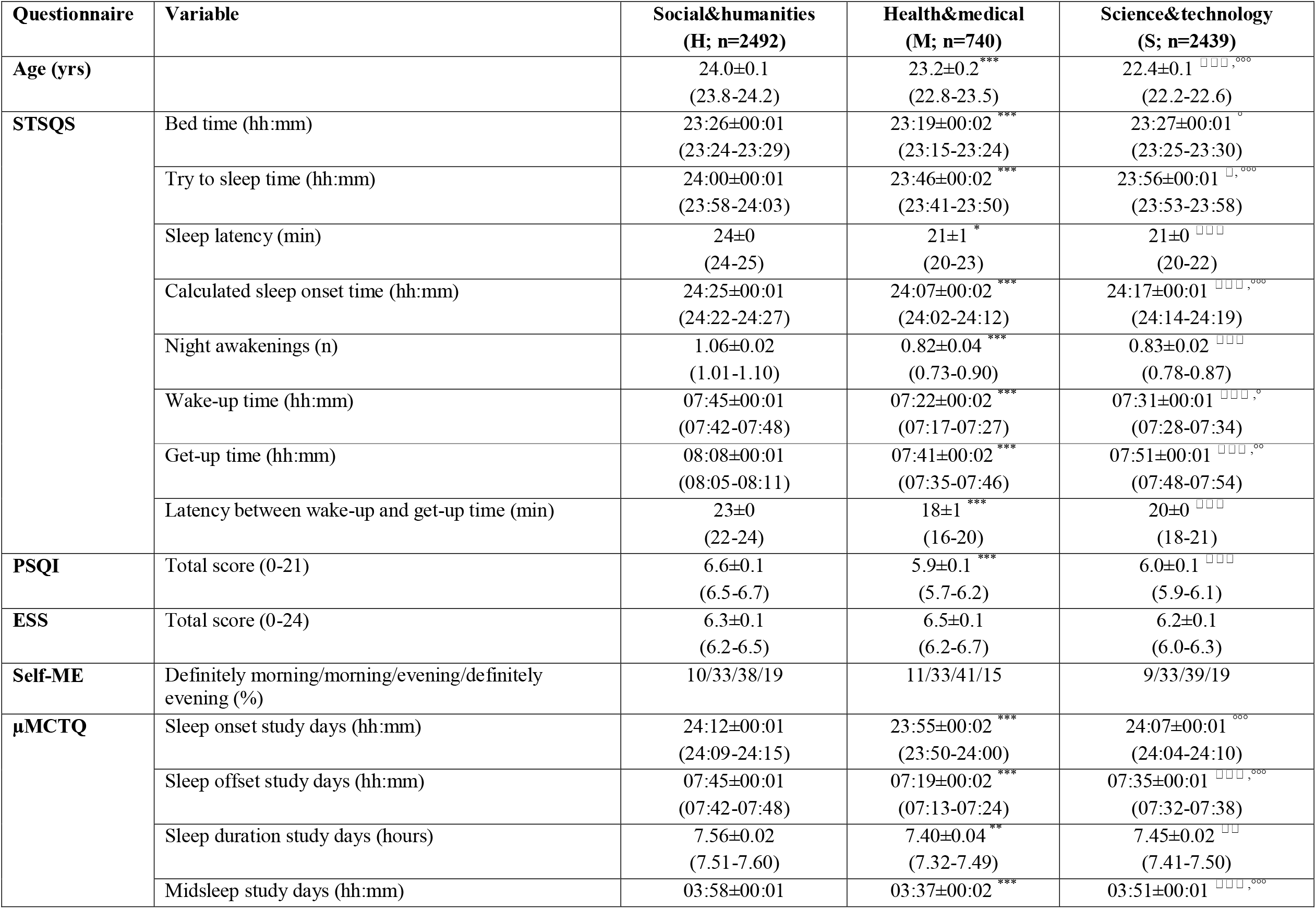
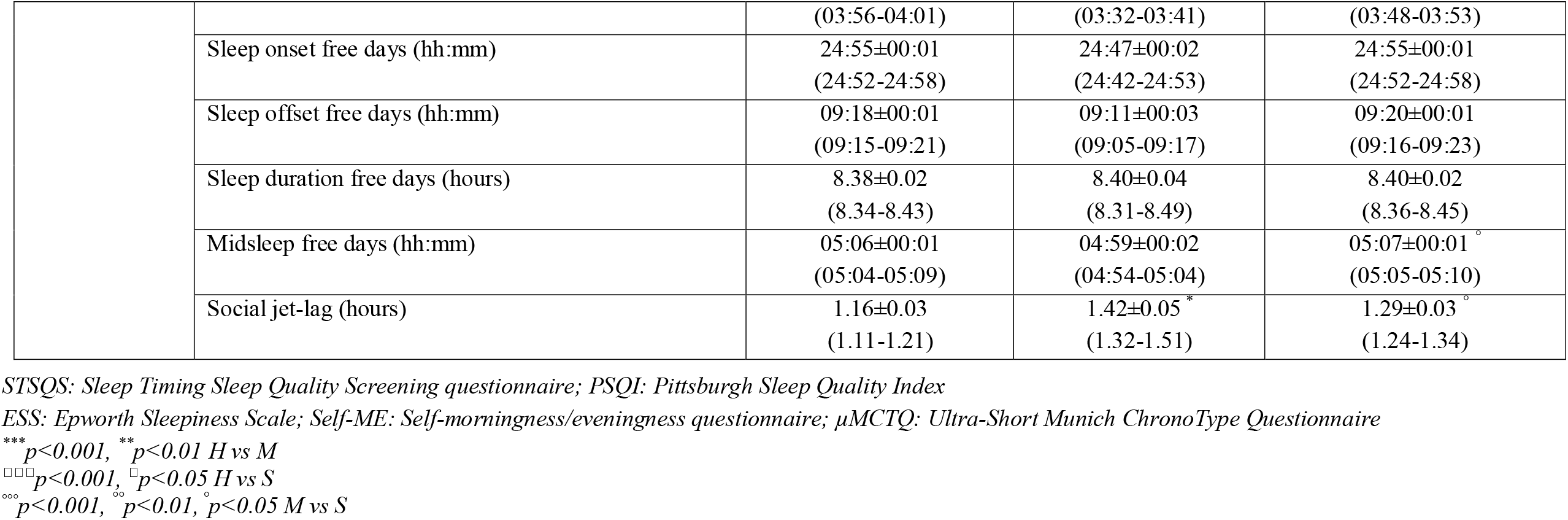
Sleep-wake indices [mean±SE (95% CI)] on joining the initiative, by study subject

Students with diseases had more night awakenings (1.1±1.2 *vs*. 0.9±1.1, p<0.0001) and worse sleep quality (PSQI: 7.0±3.5 *vs*. 6.1±2.9, p<0.0001); the same applied to students on medication.

Students who joined in October 2020 (during the pandemic) had later sleep-wake timing (e.g. wake-up time: 07:48±01:12 *vs*. 07:24±01:06, hh:mm p<0.001), less social jetlag (1.2±1.7 *vs*. 1.3±1.1 h, p<0.001), better sleep quality (PSQI abnormal 49% *vs*. 52%: χ^2^=4.6, p<0.05) and less daytime sleepiness (ESS abnormal 10% *vs*. 14%: χ^2^=14.9, p<0.001) compared to the pre-pandemic October 2019 cohort. Of note, the two cohorts were comparable in age and female/male ratio, and any students who joined in between or later were excluded for purposes of this specific comparison.

### Circadian hygiene intervention and self-reported compliance

Questionnaire completion rates decreased over time (Supplementary Table 1). While several hundred students completed the questionnaires at any given T (Supplementary Table 1), a total of 30 completed all even study times up to T12 in the *“A regular life”* group, and 28 in the *“Bright days and dark nights”* group. The two groups were comparable for sleep quality/timing, and female/male, study subject and chronotype ratios. Significant differences between the two intervention groups were observed in get-up time and in the latency between wake-up and get-up time until T10 (earlier get-up time and shorter latency in the *“Bright days and dark nights”* group). Statistical significance was lost when T12, which coincided with the transition from DST to standard time (ST), was included (Figure 2A-B). At this moment, it was the behaviour of the *“A regular life”* group that changed the most (Figure 2A-B), with the return to ST facilitating them more than their counterparts who had already been advised to go to bed and get up earlier. Along the same lines, a trend difference (0.05<p<0.1) between the two intervention groups was observed in midsleep on study days until T10 (earlier midsleep in the *“Bright days and dark nights”* group). All other sleep quality/timing variables were comparable between the two groups (Supplementary Table 2).

**Figure 2.**
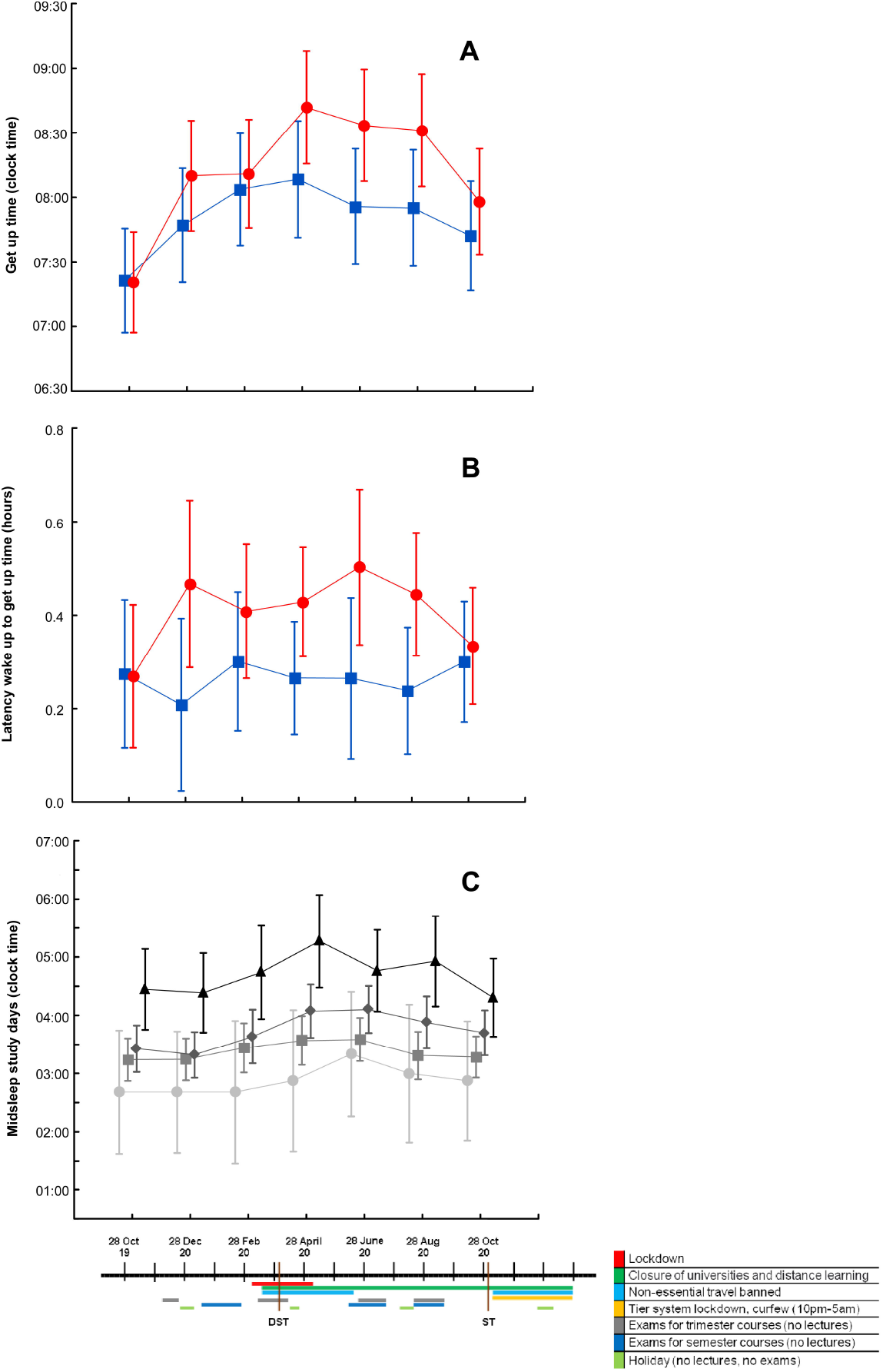
Sleep timing indices by circadian hygiene education group and by chronotype, over time and in relation to relevant 2020 events. Get-up time (**A**) and latency between wake-up and get-up time (**B**) in the *“A regular life”* (red circles; n=30 at each time point) and the *“Bright days and dark nights”* (blue squares; n=28 at each time point) groups. Differences were significant up to August 2020, i.e. T10 [get-up time: p time <0.0001 (η_p_^2^=0.189), p intervention=0.042 (η_p_^2^=0.055), p interaction n.s. (η_p_^2^ =ns); latency: p time=0.06 (η_p_^2^=ns), p intervention=0.02 (η_p_^2^=0.075), p interaction n.s. (η_p_^2^=ns)] and were abolished/reduced when October 2020 (T12), which coincided with the week after the transition from daylight saving time (DST) to standard time (ST), was included (get-up time: p time <0.0001, p intervention n.s.; p interaction n.s.; latency: p time <0.0001, p intervention=0.08; p interaction n.s.). Midsleep on study days (**C**) in definitely morning (light grey circles), morning (grey squares), evening (dark grey diamonds) and definitely evening (black triangles) chronotypes (p time <0.001, p chronotype<0.005, p interaction n.s., at both T10 and T12). Values are expressed as means ± 95% Confidence Intervals.

Significant changes in most sleep quality/timing variables were observed in both groups over time, most likely in relation to the combination of the shift to/from DST and the course of pandemic-related events that characterised 2020 (Figure 2A, 2B and 2C). This is why time points were not cumulated to increase statistical power. When students were classified into four groups based on the Self-ME, their actual sleep-wake timing reflected their self-assessment at T0. Over time, the four Self-ME groups responded in a similar fashion but to a different extent to the 2020 events (Figure 2C). When sleep-wake timing was compared between commuters and on-site students at T0 (Oct 2019) and T6 (full lockdown in April 2020), commuters’ sleep-wake habits became more similar to those of on-site students, and their social jetlag decreased (Supplementary Figure 1).

Compliance completion rates decreased over time (Supplementary Table 1); 40 students completed all odd study times in the *“A regular life”* group, and 45 in the *“Bright days and dark nights”* group. Differences between the two groups were observed in compliance to sleep-related advice, which was greater in the *“A regular life”* group (Figure 3A). Compliance to all types of advice increased over time in both groups (Figure 3A-D). Consistent differences in compliance to different types of advice were observed in both groups (meals > sleep > exercise in the *“A regular life”* group; p<0.001, all *post hoc* significant; meals and light environment > exercise > sleep in the *“Bright days and dark nights”* group; p<0.001, all *post hoc* significant except for meals *vs*. light environment). These relationships held true when analysed at T1 (highest single completion rate with 920 students in the *“A regular life”* and 841 in the *“Bright days and dark nights”* groups), at complete odd time points cumulated (280 *vs*. 315) and at all time points cumulated (3585 *vs*. 3282) (Supplementary Table 3).

**Figure 3.**
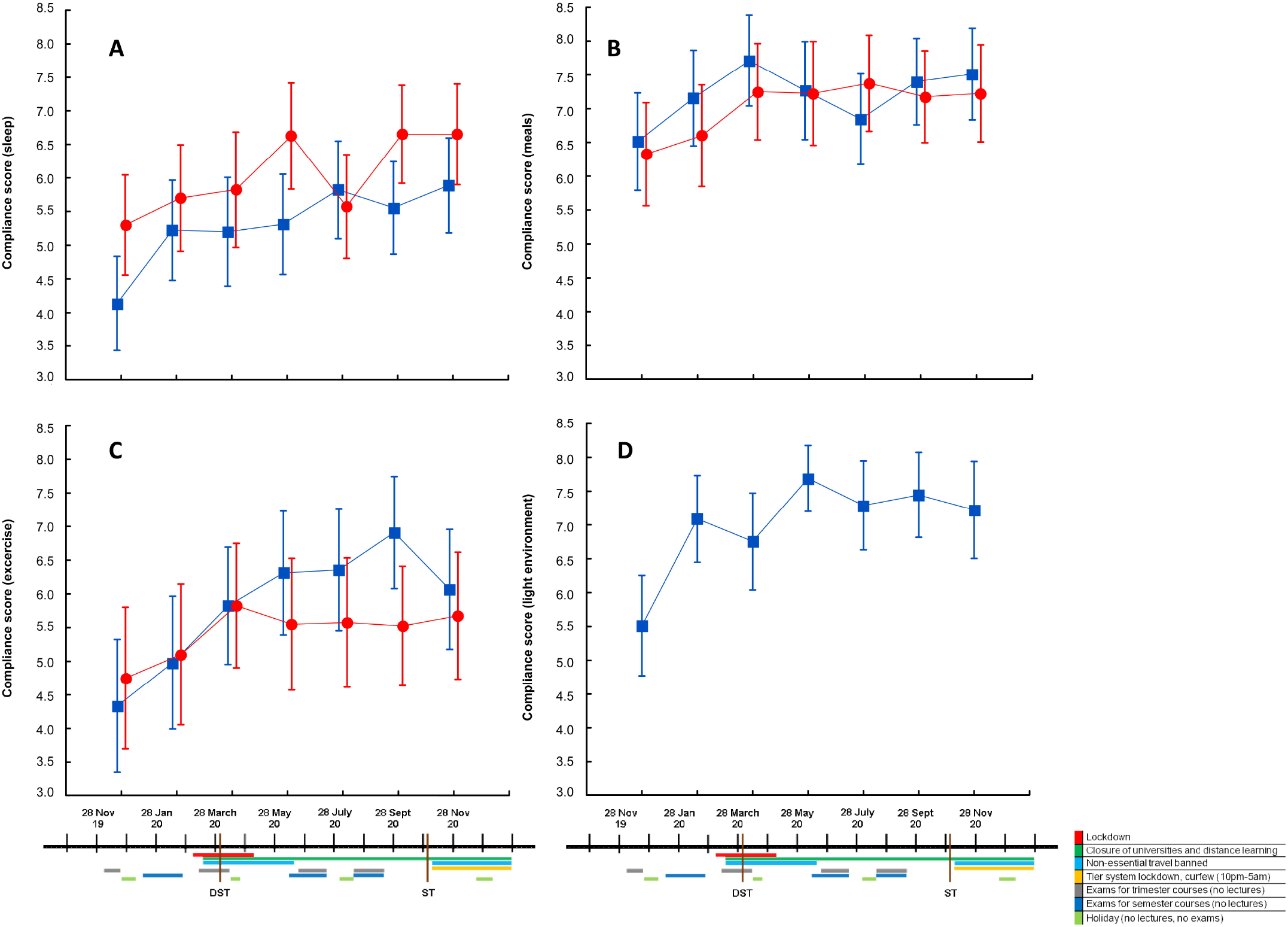
Compliance to different types of advice by circadian hygiene education group, over time and in relation to relevant 2020 events. Self-reported compliance to advice relating to sleep (**A**), meals (**B**), exercise (**C**) and light environment (**D**) in the *“A regular life”* (red circles; n=40 at each time point) and the *“Bright days and dark nights”* (blue squares; n=45 at each time point) groups. Significant differences between the two groups were observed in compliance to sleep advice (**A**), which was greater in the *“A regular life”* group (p time <0.0001, p group=0.045, p interaction n.s.). Compliance to all types of advice increased over time in both groups (p time always significant, with no further differences between groups and no interactions). Values are expressed as means ± 95% Confidence Intervals.

### Academic performance

Females had better academic performance than males at all time points. Morning had better academic performance than evening chronotypes, within each study subject (Figure 4A). Students with good sleep quality (PSQI<5) had better marks at all time points [e.g. 26.3 ± 2.4 (out of 30, 18=pass) *vs*. 25.9±2.5 on study entrance T0].

**Figure 4.**
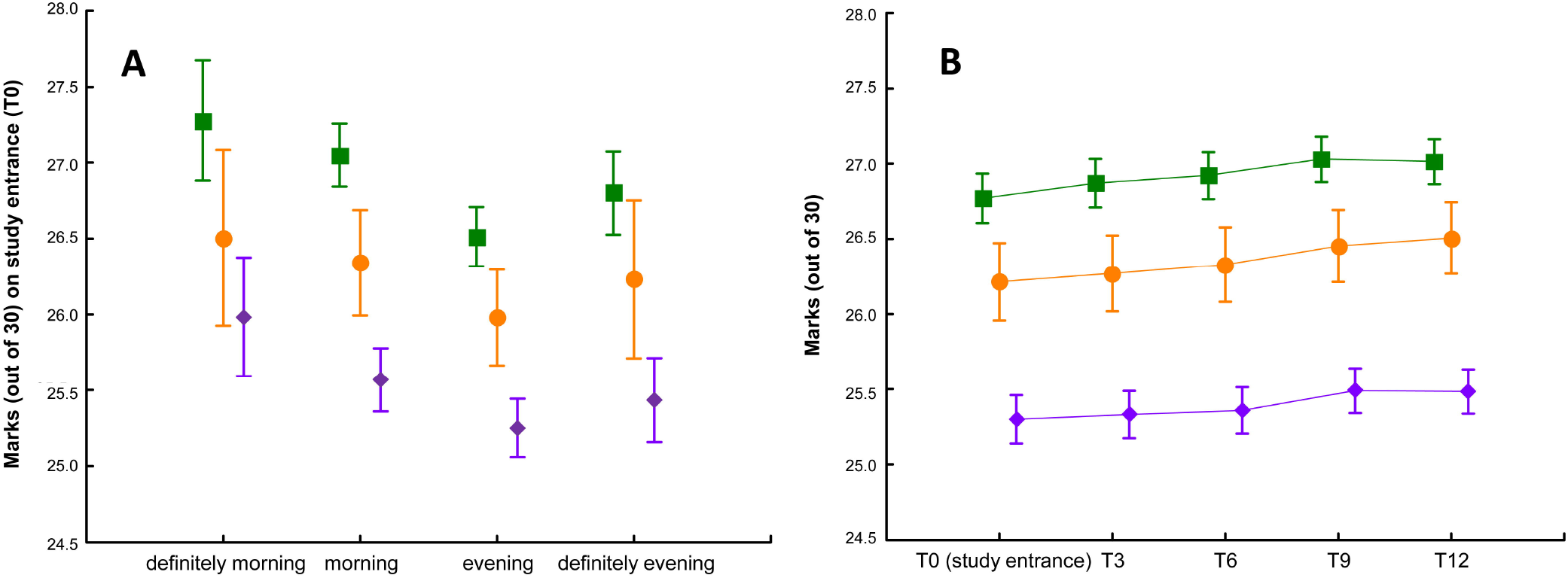
Average marks by study subject, chronotype and over time. (**A**) Average marks (out of 30) by study subject [science&technology (S): purple diamonds; health&medical (M): orange circles; social&humanities (H): green squares] and by chronotype (definitely morning n=349, morning n=1193, evening n=1390, definitely evening n=647) (p chronotype <0.0001, p study subject <0.0001; p interaction n.s.) on study entrance (T0). (**B**) Average marks (out of 30) by study subject [science&technology (S, n=858): purple diamonds; health&medical (M, n=332): orange circles; social&humanities (H, n=820): green squares] over time, aligned based on the date of joining the initiative (p time <0.0001, p study subject <0.0001; p interaction <0.0001). Values are expressed as means ± 95% Confidence Intervals.

As expected, the number of exams/credits increased over time. Average marks also increased over time (Figure 4B). Students’ academic indices were comparable between the two circadian hygiene advice groups at T0, and remained so over time.

There were institutional differences in the timing of classes for M, S and H students. Based on lectures room bookings and attendance data, M students started earlier than S students, who started earlier than H students. Further, total lecture time was longer for M compared to S/H students. Morning exams were scheduled earlier for M compared to S/H students and afternoon exams were scheduled later for H compared to M/S students (Table 3).

**Table 3.**
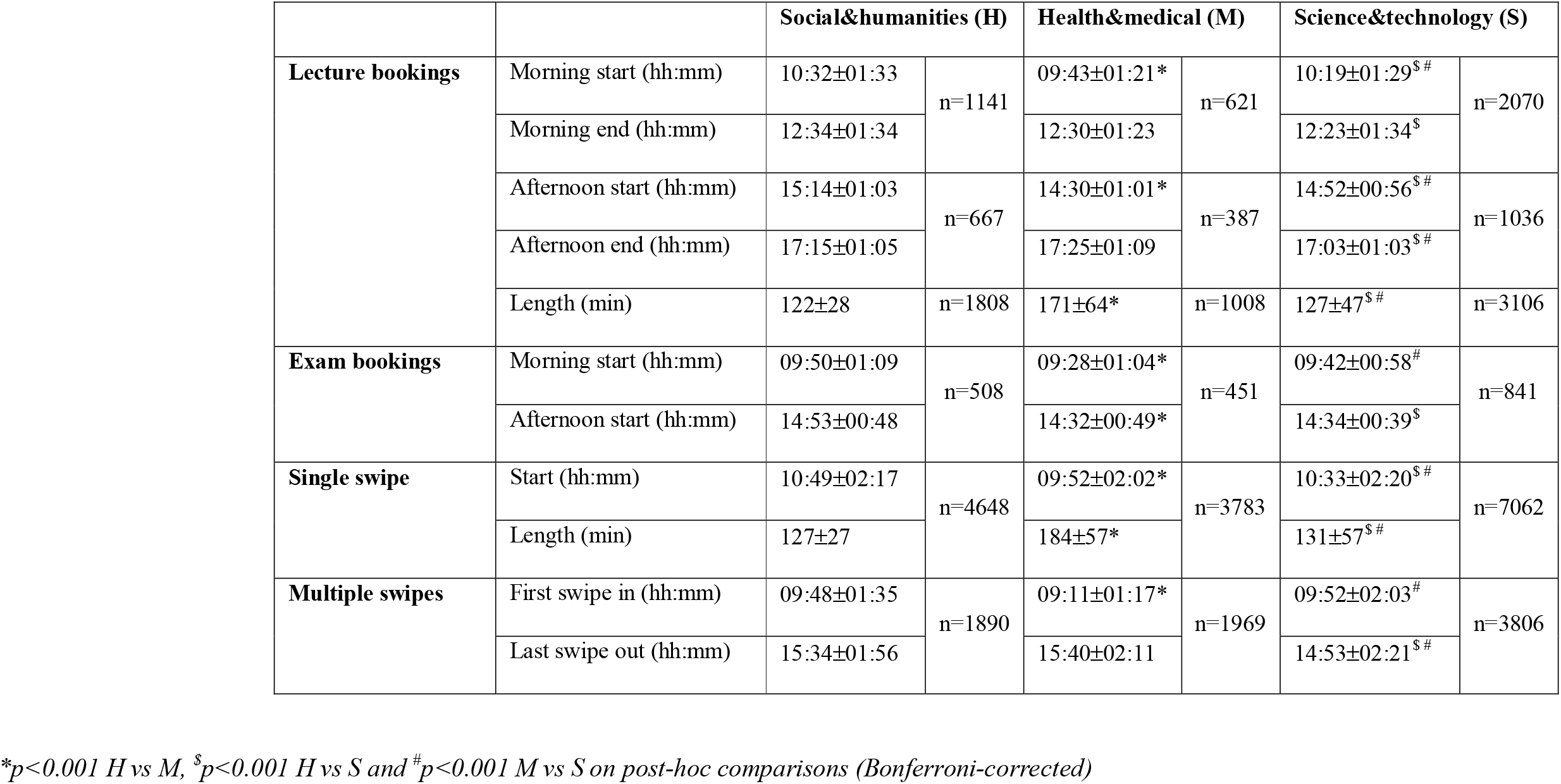
Lecture and exam booking times, plus electronic attendance records, by study subject

|  |  | Social&humanities (H) |  | Health&medical (M) |  | Science&technology (S) |  |
| --- | --- | --- | --- | --- | --- | --- | --- |
| **Lecture bookings** | Morning start (hh:mm) | 10:32±01:33 | n=1141 | 09:43±01:21* | n=621 | 10:19±01:29<sup>S #</sup> | n=2070 |
|  | Morning end (hh:mm) | 12:34±01:34 |  | 12:30±01:23 |  | 12:23±01:34<sup>S</sup> |  |
|  | Afternoon start (hh:mm) | 15:14±01:03 | n=667 | 14:30±01:01* | n=387 | 14:52±00:56<sup>S #</sup> | n=1036 |
|  | Afternoon end (hh:mm) | 17:15±01:05 |  | 17:25±01:09 |  | 17:03±01:03<sup>S #</sup> |  |
|  | Length (min) | 122±28 | n=1808 | 171±64* | n=1008 | 127±47<sup>S #</sup> | n=3106 |
| **Exam bookings** | Morning start (hh:mm) | 09:50±01:09 | n=508 | 09:28±01:04* | n=451 | 09:42±00:58<sup>#</sup> | n=841 |
|  | Afternoon start (hh:mm) | 14:53±00:48 |  | 14:32±00:49* |  | 14:34±00:39<sup>S</sup> |  |
| **Single swipe** | Start (hh:mm) | 10:49±02:17 | n=4648 | 09:52±02:02* | n=3783 | 10:33±02:20<sup>S #</sup> | n=7062 |
|  | Length (min) | 127±27 |  | 184±57* |  | 131±57<sup>S #</sup> |  |
| **Multiple swipes** | First swipe in (hh:mm) | 09:48±01:35 | n=1890 | 09:11±01:17* | n=1969 | 09:52±02:03<sup>#</sup> | n=3806 |
|  | Last swipe out (hh:mm) | 15:34±01:56 |  | 15:40±02:11 |  | 14:53±02:21<sup>S #</sup> |  |

\* $p < 0.001$ H vs M, <sup>S</sup> $p < 0.001$ H vs S and <sup>#</sup> $p < 0.001$ M vs S on post-hoc comparisons (Bonferroni-corrected)

When variables shown to affect sleep and/or academic performance [sex, study subject, commuting, sleep quality (continuous) and chronotype, plus age (continuous) as an adjustment factor] were included in a linear regression model, study subject (M>H>S), age (positive correlation), sleep quality (positive correlation) and sex (F>M) were associated with a higher number of exams passed/credits acquired, while study subject (H>M>S), chronotype (M>E) and sleep quality (positive correlation) were associated with better marks (Supplementary Table 4).

## Discussion

Poor sleep quality was common, with over half of the students reporting abnormal, albeit mild, sleep disruption. Despite reasonable sleep duration, social jetlag - a marker of sleep deprivation during study days - was also extremely common. All such abnormalities were less pronounced in the October 2020 compared to the October 2019 cohort, suggesting that long periods of full/partial lockdown and distance learning (thus less pressure from the “social clock”) made it easier for students to follow their inclination towards later sleep timing, and to sleep better. This is supported also by the large decrease in social jetlag observed in commuters between October 2019 and April 2020, and is in line with similar, albeit mostly retrospective and unpaired observations in varying populations (Korman et al., 2020; Hallman et al., 2021; Blume et al., 2020; Gao et al., 2020; Leone et al., 2020; Partinen et al., 2020; Cellini et al., 2021) and also in University students (Wright et al., 2020; Marelli et al., 2021). Most of the above studies on the effects of COVID-19-related lockdown measures and circadian/sleep variables in students and other populations were initiated during the pandemic, asking participants to provide pandemic and pre-pandemic information (the latter by recall) at a single moment in time. This methodology can be, of course, accepted but is far from ideal, because recollection of sleep-wake information (as recollection of other subjective variables) has been associated with both recall bias (Sukul et al., 2020) and summary distortion (Montagnese et al, 2009). By contrast, we were able to acquire repeat circadian/sleep information prior to and during the pandemic in a clean, longitudinal fashion, and with high time resolution.

The *“Bright days and dark nights”* advice resulted in earlier get-up time/earlier midsleep and decreased latency between wake-up and get-up time, with no impact on sleep duration. These are desirable, relevant effects in young adults (Bjorvatn et al., 2009), and were observed within the context of “a low intensity” intervention to promote healthy circadian behaviours in university students. One must remember that expectations on the real life effects of educational interventions in healthy individuals are low, as unsolicited advice is rarely complied with, there is an inherent resistance to changing habits (dietary advice is the most common reference in this respect; Kapur et al., 2008; Martis et al., 2018) and this is even more pronounced in healthy individuals, who have limited incentive to do so. Also, while we opted for a comprehensive baseline sleep-wake assessment, the intervention was circadian in its essence, and meant to affect sleep timing rather than sleep quality variables, which is what happened. The effect on sleep timing was observed despite students reporting difficulties in complying with sleep advice, and may be due, in part, to them finding it easier to follow light environment advice. This type of light advice, in a population that makes large use of portable devices (Gringas et al., 2015) in the evening/early night hours for study and recreational purposes, may indeed be crucial, also explaining part of the differences in the effects of the *“Bright days and dark nights”* and the *“A regular life”* advice. Being a University-based educational initiative, the study obviously lacks a placebo arm, and some of the methodological rigour associated with pre-ordinated design. On the positive side though, the ecological nature of the presented results provides a phenomenal amount of real-life information, which may be useful to plan similar initiatives or more formal studies in future. The initial response to the initiative, as measured by standard e-mail marketing indices, compares very favourably with average responses to Italian work/education-related initiatives in which candidate participants were contacted by email [https://www.mailup.com/; unique e-mail opens: 51.6 (our initiative) vs. 10.0%; unique link clicks: 2.9 vs. 0.5%; unique link clicks per unique e-mail opens: 5.7 vs. 4.8%]. Completion of questionnaires/compliance decreased considerably over time, suggesting feasibility issues with the initiative recall/refresh frequency, which may need modulating. It should be highlighted, however, that our intervention, lasting one year and based on monthly monitoring, was much longer and included considerably more time points than the few published similar interventions (Hershen and O’Brien, 2018; Illingworth et al., 2019; van Rijn et al., 2020; Semsarian et al., 2021). For example, Semsarian and co-authors (Semsarian et al., 2021) report 35% participation on second contact; our was similar, with 32% of students responding at T1 (Supplementary Table 1). In addition, lower completion rates after T0 do not necessarily imply lack of compliance with the advice received on joining the initiative. In addition, should more or less compliant students adhere to some or all the received *“Bright days and dark nights”* advice over long periods (for example, it is very easy to imagine them making a habit of using the night-shift function on their portable devices), its positive effects may cumulate over time. Selection bias in relation to varying compliance levels is possible but unlikely, as students who completed all procedures at all times did not have distinctive demographic/sleep features. Finally, while it is possible to hypothesise that the *“Bright days and dark nights”* advice provided to the small number of students with a definitely morning chronotype and/or a negative social jetlag may have been counterproductive, it should be highlighted that even definitely morning students had a social jet-lag which was close to one hour, thus most likely benefitting from the suggestions received.

The published, original and also review/meta-analytic literature on education (mostly sleep but also circadian) aimed at adolescents (Chung et al., 2017; Illingworth et al., 2019; van Rijn et al., 2020) or university students (Dietrich et al., 2016; Hershen and O’Brien, 2018; Semsarian et al., 2021) is not conclusive, it is based on shorter programmes, yielding generally short-lived effects. In addition, it suggests that more consistent interventions compared to ours in terms of provision of information on the science/medical evidence behind the advice delivered tend to result in increased knowledge but not necessarily in significant changes in sleep/circadian habits (Illingworth et al., 2019; van Rijn et al., 2020). Along the same lines, and despite being very appreciative of the importance of entrainment dynamics [synchronisation between the endogenous and the solar clock (Roenneberg and Merrow, 2007)], our intervention was aimed at reducing the time gap between the students’ endogenous clock and the social one, because for the time being lesson/exam times, like most social constraints, cannot be changed in the real world. So while the advice provided might not have necessarily favoured/paralleled entrainment in natural conditions, it will have hopefully helped the students functioning in their real world, with all its imposed time constraints, including lesson/exam times and DST.

The time course of sleep timing/quality between October 2019 and October 2020 suggests that the mixture of full/partial lockdown, distance learning and the transition to/from DST all had profound and intertwining effects, with any form of relaxation of the “social clock” resulting in delayed sleep timing, longer sleep duration and better sleep quality. These changes were always more obvious in evening types. Evening types also had worse academic performance compared to their more morning counterparts, within any study subject area. This is only partially in line with previous observations in high school students, whose performance was affected by chronotype mostly for scientific subjects (Zerbini et al., 2017).

Average marks increased over time, albeit very slightly. This might be explained by the fact that there is a selection process throughout university, with specific subjects – which may be more interesting for the students – being usually concentrated towards the end of their careers. However, it should be highlighted that the Italian system does not allow for a significant amount of choice/changes of the core curriculum, and these choices are probably of limited impact over the course of a single year, which was our analysis period. Alternative possibilities include some form of progressive familiarisation with the exam system, especially for younger students, or even distance learning during 2020 affecting both students’ preparation and professors’ attitudes.

In the current study, both sleep quality and chronotype independently predicted academic performance, together with study subject. Based on these data and the observed, institutional differences in lecture timing/duration and exams timing between study subjects, two approaches could be envisaged: *i)* delaying science&technology timetables by 30-60 minutes (overall study day duration would allow to accommodate this as the last swipe out for the science&technology students was between 30 and 45 minutes earlier compared to the other two study subjects; Table 3); *ii)* acquiring chronotype information from students (by the one-question Self-ME, which is fast and convenient), and taking these into account when designing timetables, with a view to limit pressure on evening chronotypes. This not only seems feasible and worth investigating but may also become necessary in the post-pandemic era, which will presumably be still characterised by the need to avoid large groups of students attending lectures at any given time. Thus should lectures/exams timetable shifts remain necessary in large universities such as ours, they should probably be better designed to maximise academic performance. Similar approaches (i.e. acquisition and use of chronotype information to design work shifts) have already proven effective in terms of sleep length in workers from major industries (Vetter et al., 2015). In addition, the advice shown to be easier to follow (i.e. advice on meal timing and light environment from *“Bright days and dark nights”*) could readily be provided to students when they register at the University as part of their induction package.

In conclusion, this large and comprehensive set of baseline data, the response and compliance to the educational initiative, the time course of sleep timing in the year of the pandemic, and the observed differences in academic performance by chronotype all underlie the importance of designing circadian-friendly university timetables.

## Data Availability

All data produced in the present study are available upon reasonable request to the authors

## ACKNOWLEDGEMENTS

The authors are grateful to Gioia Grigolin, Rosa Nardelli, Valentino Callegari, and Elena Carnevali, at the University of Padova, for their invaluable help with the technical and logistic aspects of the initiative.

