## Supplementary Tables and Figures for "Sleep and circadian indices for planning post-pandemic university timetables"

**Supplementary Table 1.** Study design, information acquisition and circadian hygiene advice provision timing, with completion rates

| **Time from study entrance (months)** | **Sleep-wake questionnaires** | **Compliance** | **Advice** |
| --- | --- | --- | --- |
| **T0 (even)** | √ (n=5740) |  | √ |
| **T1 (odd)** |  | √ (n=1853) | √ |
| **T2 (even)** | √ (n=842) | √ (n=763) | √ |
| **T3 (odd)** |  | √ (n=444) | √ |
| **T4 (even)** | √ (n=491) | √ (n=258) | √ |
| **T5 (odd)** |  | √ (n=153) | √ |
| **T6 (even)** | √ (n=430) | √ (n=112) | √ |
| **T7 (odd)** |  | √ (n=98) | √ |
| **T8 (even)** | √ (n=265) | √ (n=76) | √ |
| **T9 (odd)** |  | √ (n=78) | √ |
| **T10 (even)** | √ (n=259) | √ (n=69) | √ |
| **T11 (odd)** |  | √ (n=50) | √ |
| **T12 (even)** | √ (n=222) | √ (n=43) | √ |
| **T odd n** |  | √ | √ |
| **T even n** | √ | √ | √ |

**Supplementary Table 2.** Sleep-wake indices [mean±SE (95% CI)] over time, by treatment group

| **Questionnaire** | **Variable** | **Group** | **28 Oct 19** | **28 Dec 19** | **28 Feb 20** | **28 April 20** | **28 June 20** | **28 Aug 20** | **28 Oct 20** |
| --- | --- | --- | --- | --- | --- | --- | --- | --- | --- |
| **STSQS** | Bed time (hh:mm) | *A regular life* | 23:00±00:10 (22:38-23:22) | 23:01±00:11  (22:39-23:24) | 23:14±00:11  (22:52-23:36) | 23:14±00:13  (22:53-23:52) | 23:29±00:11  (23:05-23:52) | 23:23±00:10  (23:01-23:45) | 23:10±00:11  (22:45-23:33) |
|  |  | *Bright days and dark nights* | 22:58±00:11  (22:36-23:21) | 23:05±00:11  (22:42-23:28) | 23:15±00:11  (22:52-23:37) | 23:29±00:14  (23:01-23:57) | 23:30±00:12  (23:05-23:54) | 23:13±00:11  (22:50-23:36) | 23:09±00:11  (22:45-23:33) |
|  | Try to sleep time (hh:mm) | *A regular life* | 23:37±00:10  (23:15-23:59) | 23:40±00:10  (23:19-24:00) | 23:31±00:10  (23:09-23:52) | 24:00±00:12  (23:35-24:26) | 24:00±00:10  (23:39-24:20) | 23:58±00:10  (23:36-24:19) | 23:40±00:10  (23:19-24:01) |
|  |  | *Bright days and dark nights* | 23:23±00:11  (23:00-23:45) | 23:26±00:10  (23:05-23:47) | 23:38±00:11  (23:15-24:00) | 23:54±00:13  (23:28-24:20) | 23:43±00:10  (23:22-24:05) | 23:45±00:11  (23:23-24:07) | 23:38±00:10  (23:17-24:00) |
|  | Sleep latency (min) | *A regular life* | 19±2  (14-23) | 19±2  (15-23) | 17±2  (13-22) | 18±2  (13-23) | 20±2  (15-25) | 15±2  (10-20) | 14±2  (10-18) |
|  |  | *Bright days and dark nights* | 14±2  (9-19) | 14±2  (10-19) | 13±2  (9-18) | 13±3  (8-19) | 13±3  (8-18) | 14±2  (9-19) | 14±2  (10-18) |
|  | Calculated sleep onset time (hh:mm) | *A regular life* | 23:56±00:10  (23:34-24:17) | 23:59±00:10  (23:38-24:19) | 23:48±00:10  (23:26-24:09) | 24:18±00:12  (23:53-24:44) | 24:20±00:10  (23:59-24:41) | 24:13±00:11  (23:51-24:35) | 23:54±00:10  (23:33-24:15) |
|  |  | *Bright days and dark nights* | 23:37±00:11  (23:15-23:59) | 23:40±00:10  (23:19-24:02) | 23:51±00:11  (23:28-24:13) | 24:07±00:13  (23:41-24:34) | 23:56±00:10  (23:34-24:18) | 23:59±00:11  (23:36-24:22) | 23:52±00:10  (23:30-24:14) |
|  | Night awakenings (n) | *A regular life* | 0.80±0.18  (0.45-1.15) | 0.77±0.16  (0.45-1.08) | 0.80±0.16  (0.48-1.11) | 0.70±0.15  (0.39-1.00) | 0.57±0.14  (0.29-0.85) | 0.60±0.13  (0.34-0.86) | 0.63±0.13  (0.36-0.90) |
|  |  | *Bright days and dark nights* | 0.75±0.18  (0.38-1.11) | 0.79±0.16  (0.46-1.11) | 0.79±0.16  (0.46-1.11) | 0.71±0.16  (0.40-1.03) | 0.71±0.14  (0.42-1.00) | 0.68±0.14  (0.40-0.95) | 0.73±0.14  (0.45-1.01) |
|  | Wake-up time (hh:mm) | *A regular life* | 07:04±00:11  (06:40-07:27) | 07:42±00:11  (07:19-08:04) | 07:46±00:12  (07:22-08:10) | 08:16±00:12  (07:50-08:41) | 08:03±00:12  (07:38-08:28) | 08:04±00:13  (07:38-08:30) | 07:37±00:11  (07:15-08:00) |
|  |  | *Bright days and dark nights* | 07:04±00:12  (06:40-07:29) | 07:34±00:11  (07:10-07:58) | 07:45±00:12  (07:20-08:10) | 07:52±00:13  (07:25-08:19) | 07:39±00:13  (07:13-08:05) | 07:40±00:13  (07:13-08:07) | 07:24±00:11  (07:00-07:47) |
|  | Get-up time (hh:mm) | *A regular life* | 07:20±00:11  (06:57-07:43) | 08:10±00:12  (07:44-08:35) | 08:11±00:12  (07:45-08:36) | 08:41±00:13  (08:15-09:08) | 08:33±00:12  (08:07-08:59) | 08:31±00:13  (08:05-08:57) | 07:58±00:12  (07:33-08:22) |
|  |  | *Bright days and dark nights* | 07:21±00:12  (06:57-07:45) | 07:46±00:13  (07:20-08:13) | 08:03±00:13  (07:37-08:29) | 08:08±00:13  (07:41-08:35) | 07:55±00:13  (07:28-08:22) | 07:55±00:13  (07:27-08:22) | 07:42±00:12  (07:16-08:07) |
|  | Latency between wake-up and get-up time (min) | *A regular life* | 16±4  (7-25) | 28±5  (17-38) | 24±4  (15-33) | 25±3  (18-32) | 30±4  (20-40) | 26±3  (18-34) | 20±3  (12-27) |
|  |  | *Bright days and dark nights* | 16±4  (7-25) | 12±5  (1-23) | 18±4  (9-26) | 15±3  (8-23) | 15±5  (5-26) | 14±4  (6-22) | 18±3  (10-25) |
| **PSQI** | Total score (0-21) | *A regular life* | 6.00±0.40  (5.20-6.80) | 5.53±0.43  (4.67-6.40) | 5.63±0.41  (4.80-6.46) | 5.47±0.43  (4.61-6.32) | 5.40±0.41  (4.58-6.21) | 5.20±0.53  (4.14-6.26) | 4.30±0.43  (3.44-5.16) |
|  |  | *Bright days and dark nights* | 4.82±0.41  (4.00-5.65) | 5.14±0.45  (4.25-6.04) | 4.36±0.43  (3.50-5.22) | 4.00±0.44  (3.11-4.89) | 4.57±0.42  (3.73-5.41) | 5.11±0.55  (4.01-6.20) | 4.71±0.44  (3.83-5.60) |
| **ESS** | Total score (0-24) | *A regular life* | 6.03±0.64  (4.74-7.32) | 5.93±0.68  (4.57-7.29) | 5.77±0.57  (4.62-6.91) | 5.14±0.62  (3.89-6.87) | 5.40±0.61  (4.18-6.62) | 5.20±0.62  (3.96-6.44) | 5.10±0.66  (3.78-6.42) |
|  |  | *Bright days and dark nights* | 5.82±0.67  (4.49-7.16) | 5.14±0.70  (3.74-6.55) | 4.86±0.59  (3.67-6.04) | 4.41±0.64  (2.86-5.42) | 4.14±0.63  (2.88-5.40) | 4.54±0.64  (3.26-5.82) | 4.50±0.68  (3.13-5.86) |
| **µMCTQ** | Midsleep study days (hh:mm) | *A regular life* | 03:36±00:10  (03:15-03:58) | 03:31±00:10  (03:10-03:53) | 03:44±00:12  (03:18-04:09) | 03:57±00:13  (03:31-04:24) | 04:07±00:10  (03:45-04:28) | 03:50±00:12  (03:25-04:15) | 03:35±00:10  (03:14-03:56) |
|  |  | *Bright days and dark nights* | 03:13±00:11  (02:51-03:36) | 03:13±00:11  (02:51-03:36) | 03:31±00:13  (03:05-03:57) | 03:53±00:13  (03:26-04:21) | 03:41±00:11  (03:18-04:04) | 03:33±00:13  (03:07-04:00) | 03:29±00:10  (03:07-03:51) |
|  | Midsleep free days (hh:mm) | *A regular life* | 04:50±00:11  (04:28-05:12) | 04:47±00:10  (04:25-05:09) | 04:39±00:11  (04:16-05:02) | 04:58±00:11  (04:36-05:21) | 04:50±00:10  (04:28-05:11) | 04:46±00:10  (04:24-05:08) | 04:37±00:10  (04:17-04:58) |
|  |  | *Bright days and dark nights* | 04:30±00:11  (04:07-04:53) | 04:35±00:11  (04:12-04:58) | 04:28±00:11  (04:05-04:52) | 04:40±00:11  (04:16-05:03) | 04:38±00:11  (04:15-05:00) | 04:28±00:11  (04:05-04:51) | 04:22±00:10  (04:01-04:44) |
|  | Social jet-lag (hours) | *A regular life* | 1.25±0.15  (0.94-1.56) | 1.31±0.16  (0.98-1.63) | 0.96±0.15  (0.66-1.25) | 1.02±0.13  (0.75-1.29) | 0.75±0.10  (0.56-0.95) | 0.96±0.13  (0.69-1.23) | 1.06±0.16  (0.74-1.37) |
|  |  | *Bright days and dark nights* | 1.34±0.16  (1.01-1.66) | 1.38±0.17  (1.03-1.72) | 0.94±0.15  (0.63-1.25) | 0.83±0.14  (0.55-1.12) | 0.98±0.10  (0.77-1.18) | 0.93±0.14  (0.64-1.22) | 0.90±0.17  (0.56-1.23) |

**Supplementary Figure 1.** Social jetlag in on-site students (black squares; n=140) and commuters (grey circles; n=77) in October 2019 and in April 2020 (full lockdown); p group n.s.; p time <0.0001; p interaction <0.01. Values are expressed as means ± 95% Confidence Intervals.

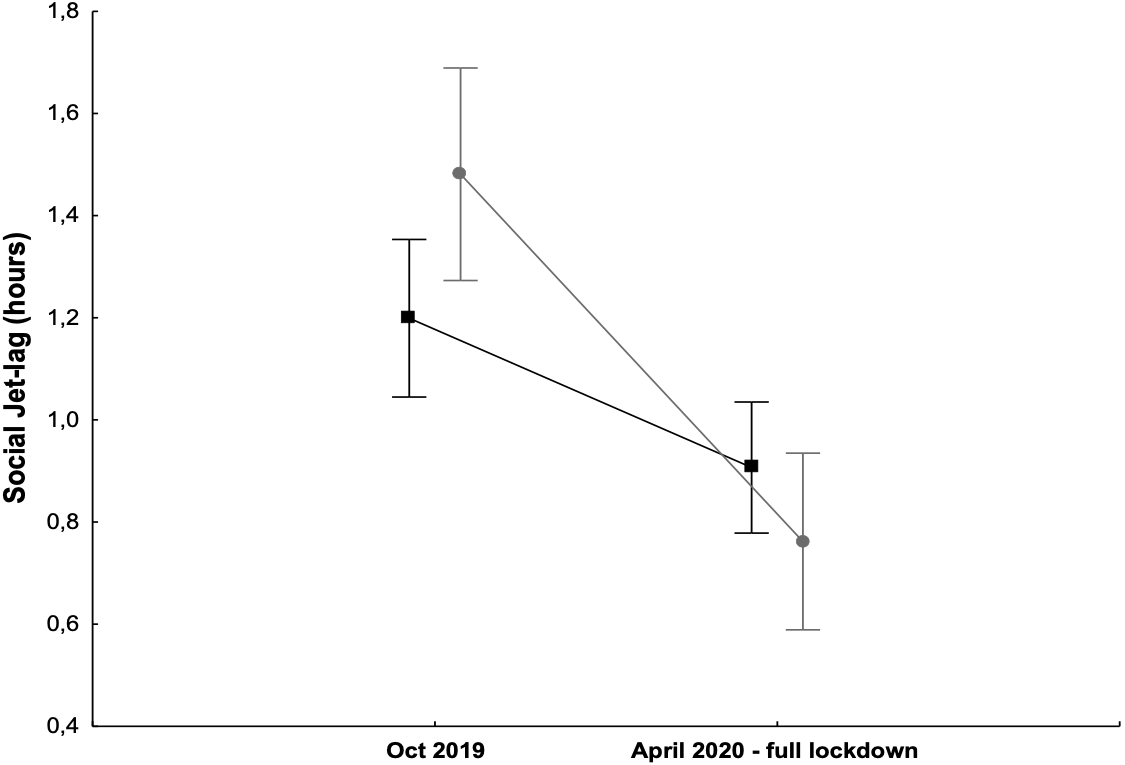

1.8

1.6

1.2

1.4

1.0

0.8

0.6

0.4

**Supplementary Table 3.** Average self-reported compliance values (on a 0-10 visual-analogue scale), by advice group, at T1 (November 2019), at all complete odd times cumulated and at all times cumulated.

|  |  | ***“A regular life”*** | | ***“Bright days and dark nights”*** | |
| --- | --- | --- | --- | --- | --- |
|  |  | **n** | **Mean±SD** | **n** | **Mean±SD** |
| **T1 Nov 2019** | **Sleep** | 920 | 5.5±2.3 | 841 | 4.3±2.5 |
|  | **Meal** | 920 | 6.2±2.3 | 841 | 6.0±2.7 |
|  | **Exercise** | 920 | 5.1±3.1 | 841 | 4.7±3.4 |
|  | **Light** |  |  | 841 | 5.8±2.6 |
| **All complete odd times cumulated** | **Sleep** | 280 | 6.0±2.4 | 315 | 5.3±2.6 |
|  | **Meal** | 280 | 7.0±2.2 | 315 | 7.2±2.4 |
|  | **Exercise** | 280 | 5.4±3.0 | 315 | 5.8±3.2 |
|  | **Light** |  |  | 315 | 7.0±2.3 |
| **All times cumulated** | **Sleep** | 3585 | 6.0±2.3 | 3282 | 4.7±2.6 |
|  | **Meal** | 3585 | 6.8±2.2 | 3282 | 6.3±2.7 |
|  | **Exercise** | 3585 | 5.3±3.0 | 3282 | 5.3±3.3 |
|  | **Light** |  |  | 3282 | 6.3±2.5 |

**Supplementary Table 4.** Significance and weights (the larger partial eta-squared, the larger the weight of the variable, i.e. the proportion of the variability in the dependent variables explained by the effect) of different predictors on three indices of academic performance at T0 (n=3726 for exams and credits, n=3580 for marks).

|  | **Variables** | | | | | | | | | | | |
| --- | --- | --- | --- | --- | --- | --- | --- | --- | --- | --- | --- | --- |
|  | **Age** | | **Chronotype** | | **Commuting**  **(commuters vs. on-site)** | | **Sex** | | **Sleep quality**  **(total PSQI score)** | | **Study area** | |
| **Number exams** | p<0.0001 | η_p_^2^=0.037 | p=0.255 | η_p_^2^=0.001 | p=0.397 | η_p_^2^=0.000 | p<0.0001 | η_p_^2^=0.005 | p=0.005 | η_p_^2^=0.003 | p<0.0001 | η_p_^2^=0.167 |
| **Number credits** | p<0.0001 | η_p_^2^=0.026 | p=0.311 | η_p_^2^=0.001 | p=0.413 | η_p_^2^=0.000 | p=0.009 | η_p_^2^=0.002 | p=0.007 | η_p_^2^=0.002 | p<0.0001 | η_p_^2^=0.145 |
| **Average marks** | p=0.165 | η_p_^2^=0.000 | p<0.0001 | η_p_^2^=0.008 | p=0.550 | η_p_^2^=0.000 | p=0.201 | η_p_^2^=0.000 | p<0.0001 | η_p_^2^=0.007 | p<0.0001 | η_p_^2^=0.063 |

*PSQI: Pittsburgh Sleep Quality Index*
